# COVID-19 in Hospitalized Ethiopian Children: Characteristics and Outcome Profile

**DOI:** 10.1101/2020.10.30.20223115

**Authors:** Tigist Workneh Leulseged, Ishmael Shemsedin Hassen, Endalkachew Hailu Maru, Wuletaw Chane Zewde, Negat Woldehawariat Chamiso, Mesay Gemechu Edo, Daniel Simeneh Abebe, Muktar Awol Aliy

**Author notes:** Corresponding author: Tigist Workneh Leulseged, Research development office, Millennium COVID-19 Care Center, Addis Ababa, Ethiopia and Department of Internal Medicine, St. Paul’s Hospital Millennium Medical College, Addis Ababa, Ethiopia.

## Abstract

**Background:** Considering the number of people affected and the burden to the health care system due to the Coronavirus pandemic, there is still a gap in understanding the disease better leaving a space for new evidence to be filled by researchers. This scarcity of evidence is observed especially among children with the virus. Therefore, this study aimed to assess the characteristics and outcome profile of children with COVID-19 admitted to Millennium COVID-19 Care Center in Ethiopia.

**Methods:** A prospective cohort study was conducted among 90 children with COVID-19 who were admitted from June 23 to September 17, 2020. Data was summarized using frequency tables, mean ± standard deviation or median with Inter Quartile range values. A chi-square test/ Fischer’s exact test was used to compare disease severity between groups.

**Results:** The median age of the participants was 15 years and 57 were females. The most common reported route of disease transmission was through close contact with a diagnosed person (41/90). Only three had a history of pre-existing comorbid illness. One-third (31/90) had one or more symptoms at diagnosis, the most common being cough (20/90). Among the 90 patients, 59 were asymptomatic, 14 had mild disease and the rest 17 had moderate disease. Based on the chi-square/ Fischer’s exact test result, no statistically significant difference was observed between the age groups and sex.

**Conclusions:** Pediatric patients seemed to have a milder disease presentation and a favorable outcome compared to other countries report and also the adult pattern observed in our country.

## BACKGROUND

According to the World Health Organization weekly epidemiological update the Corona Virus pandemic has affected more than 83, 326, 479 individuals and resulted in 1,831, 703 deaths worldwide, as of January 3, 2020 [1]. Age classified report of the disease in terms of incidence, severity and outcome showed that the pandemic affects all age groups and that younger age groups have far less probability of acquiring the disease and with milder disease course and favorable outcome if they do get infected [2].

Studies conducted till now have focused mainly on adult patients, resulting in less understating of the disease presentation and outcome among children especially in the African set up According to studies conducted in UK, US and Canada, the disease seemed to be common among males just like the report from adults [3, 4] andthe commonly affected age groups are in the two extremes, infants and adolescent [3, 5]. The majority of children are reported to acquire the disease through household contact with adults [6, 7].

Both symptomatic and asymptomatic cases are reported, with a number of studies including systematic review report showing that a higher proportion of children had one or more symptom at presentation. The commonly reported symptoms are fever and cough, with fever being the predominant symptom in many countries. Nonspecific symptoms like fatigue and myalgia are also reported [6-9]. This is unlike the adult reports where respiratory symptoms were recorded to be the commonest [10-16].

Mild and moderate diseases are commonly reported with severe and critical cases observed in relatively few cases [6, 8, 9]. Admission to critical care was determined by age and race [3]. Underlying comorbid illness was found in a considerable proportion of hospitalized patients ranging from 42% in UK to 83% in US and Canada [3, 4].

A study conducted in South Africa among children younger than 13 years showed that among hospitalized children, majority were symptomatic, one-fifth had pre-existing medical condition and respiratory support was required in half of the children [17].

Laboratory derangements including lymphopenia, leucopenia, increased D-dimer and increased Creatinine are reported as common findings. Chest x-ray reports of children with pneumonia showed that more than half had a ground glass opacity pattern [8, 9].

Possible outcomes reported range from a complete uneventful recovery to complications like multiple organ failure and multiple inflammatory syndrome up to death. A more favorable disease outcome is observed among children compared to adults with few reports of complications and mortality rate of 1-4% among hospitalized children [3-6].

Due to scarcity of study among Children with COVID-19 in Africa, understanding the disease pattern and its outcome is crucial to guide decision making. Therefore, in this study we aimed to assess the characteristics and outcome profile of children with RT-PCR confirmed COVID-19 and admitted to Millennium COVID-19 Care Center in Ethiopia.

## METHODS

### Study Setting, Design and Population

An institution based prospective cohort study design was conducted at Millennium COVID-19 Care Center (MCCC), a makeshift hospital in Addis Ababa, Ethiopia. The center is remodeled from the previous Millennium hall which was a recreational center. The center started accepting patients on June 2, 2020 with a Capacity of 1000 Beds, 40 ICU beds and 12 mechanical ventilators. At that time the national admission criteria was having positive RT-PCR test result irrespective of the presence of symptoms or severity of the disease including cases identified through contact tracing. Therefore, the Center was used as both quarantine and treatment center in order to halt the transmission of the disease.

The center was modelled to admit adult and none pregnant patients only but since the other centers were not ready for COVID-19 case admission, all age groups and pregnants were also being admitted for few months. The Center accepts patients from referral centers (health center up to tertiary hospital which doesn’t have COVID-19 Care Setup) only with a positive RT-PCR result and the service is provided for free.

The source population was all consecutively admitted children with COVID-19 admitted to MCCC with a confirmed diagnosis of COVID-19 using RT-PCR, as reported by a laboratory given mandate to test such patients by the Ethiopian Federal Ministry of Health and who were on follow up and/or treatment from June 23 to September 17, 2020 [18]. All eligible cases admitted during this period was prospectively observed from admission to discharge for their clinical presentation and disease outcome The follow up period is selected considering the first pediatric case admission and discharge of the last pediatric case from the center. After that period other institutions that provide COVID-19 Care service specialized for pediatric and obstetric cases were organized and our center was dedicated for management of adult cases only. Also since most pediatric cases are asymptomatic or mild, there are few pediatric admissions at the national level in general after the national admission criteria shifted to accommodate more severe cases, those with co-morbidity and older age groups.

### Sample size Determination and Sampling Technique

All consecutively admitted children with COVID-19 during the three months follow up period were included in the study. During this interval a total of 115 children with COVID-19 were admitted to the Center. Finally 90 children who fulfilled the eligibility criteria were included in the final analysis.

### Eligibility criteria

All children 18 years and younger with RT-PCR confirmed COVID-19 who were on follow up and/or treatment at the center from June 23 to September 17, 2020 and with complete follow up data were included in the study.

### Operational Definitions

#### Asymptomatic patient

any patient who tested positive for COVID-19 but does not have any symptoms. These patients are detected after isolation and contact tracing as done by the Ethiopian Public Health Institute (EPHI) ^28^.

**COVID-19 severity score**

was determined based on the WHO classification as follows ^29^.

– **Mild Disease:** Characterized by fever, malaise, cough, upper respiratory symptoms, and/or less common features of COVID-19 (headache, loss of taste or smell etc.)
– **Moderate Disease:** Patients with lower respiratory symptom/s. They may have infiltrates on chest X-ray. These patients are able to maintain oxygenation on room air.

### Data collection and quality assurance

A pre-tested interviewer administered questionnaire was used to collect data from patients and their medical charts to capture their socio-demographic, clinical and disease outcome profile.

The data was collected by trained data collectors and all the necessary precautions in order to prevent the acquisition and disseminate the virus were taken during the data collection process. To improve data quality, data cleaning through checking for inconsistencies, numerical errors and missing parameters was done. Where discrepancies are observed, data entered was verified with the primary data source.

### Statistical analysis

Once the collected data was coded and cleaned, it was exported and stored into SPSS version 23 for analysis. Categorical covariates were summarized using frequencies and percentages and numerical variables were summarized with a mean ± standard deviation (SD) or median with Inter Quartile range (IQR) values.

A chi-square test (or Fischer’s exact test for those variables which do not meet the chi-square assumptions) were used to determine the presence of a significant difference between the age groups and sex based on COVID-19 disease severity. A statistically significant difference was detected for variables with a P-value of ≤ 0.05.

### Ethics

The study was conducted after obtaining ethical clearance from St. Paul’s Hospital Millennium Medical College Institutional Review Board. Written informed consent was obtained from the participants and/or their care givers. The study had no risk/negative consequence on those who participated in the study. Medical record numbers were used for data collection and personal identifiers were not used in the research report. Access to the collected information was limited to the principal investigator and confidentiality was maintained throughout the project.

## RESULT

### Socio–demographic, co-morbid illness and presenting symptom characteristics

The median age of the participants was 15 (IQR, 10-17) years. The majority (33/90) of the participants were 15 to 18 years. Almost two third (57/90) of the participants were females. The majority (80/90) were from Addis Ababa, the capital city of Ethiopia. The most common reported route of disease transmission was through close contact with a diagnosed person (41/90).

Only three had a history of pre-existing comorbid illness, two had bronchial asthma and one had a history of valvular heart disease. One-third (31/90) of the patients had one or more symptom at admission and rest 59 were asymptomatic. The most common symptom was cough (20/90), followed by sore throat and headache (each constitutes 9 cases), fever, runny nose, chest pain and fatigue (each constitutes 5 cases) and then nausea and vomiting (4 cases). **(Table 1)**

**Table 1:** Socio–demographic and presenting symptom related variables among children with COVID-19 (n=90)

| **Variables** | **Total** |
| --- | --- |
| **Age group (in years)** |  |
| ≤ 5 | 10 |
| 6-10 | 16 |
| 11-15 | 31 |
| 15-18 | 33 |
| **Sex** |  |
| Female | 57 |
| Male | 33 |
| **Pre-existing Co-morbid illness** | 3 |
| **Asymptomatic** | 66 |
| **Cough** | 20 |
| **Sorethroat** | 9 |
| **Headache** | 9 |
| **Fever** | 5 |
| **Chest pain** | 5 |
| **Fatigue** | 5 |
| **Runny nose** | 5 |
| **Nausea/ vomiting** | 4 |

### Disease severity and outcome

Among the 90 patients, 59 were asymptomatic, 14 had mild disease and the rest 17 had moderate disease. From the 90 children, 67 were discharged improved and the rest 23 were transferred to another hospital. Among the 23 transferred cases, 17 were asymptomatic, 4 had mild disease and 2 had moderate disease. One child with valvular heart disease was transferred to a center where there is a pediatrician for better management and follow up as MCCC is setup for an adult care. Eight were transferred to another center with family request for convenience of visit. The rest 14 were transferred to another center as an adult family member with whom the children were admitted with were transferred due to different reasons.

### Management of the admitted children

Mild cases were managed with analgesic and antipyretic to relieve symptoms and the moderate cases received antibiotics treatment in addition. Since there was no severe case during the observation period, none of the children required an oxygen therapy or intensive care unit admission.

### Characteristics of the children with moderate COVID-19

Seventeen of the children had moderate COVID-19 at admission. The profile of the 17 patients showed that, 11/17 were 15-18 years old and 12/17 were females. Only one child had Valvular heart disease, the rest didn’t have any known pre-existing co-morbid illness. 15/17 were discharged improved and 2/17 were transferred to another hospital (including the child with valvular heart disease who was transferred for better care to a center with a pediatrician).

The complete blood count, renal function and liver enzyme laboratory report of the moderate cases showed that all values were with in normal range. (**Table 2**)

**Table 2:** Baseline Laboratory markers of children with moderate COVID-19 (n=17)

| **Variables** | **Mean** | **Standard deviation** |
| --- | --- | --- |
| **Complete blood cell count** |  |  |
| Hemoglobin (mg/dl) | 14.94 | 1.82 |
| Hematocrit (%) | 41.65 | 9.92 |
| White blood cell count ($\times 10^3$ Cells/L) | 6.87 | 2.33 |
| Platelet count ($\times 10^3$ Cells/ L) | 282.0 | 82.37 |
| **Renal function test** |  |  |
| Urea (mg/dl) | 27.82 | 9.88 |
| Creatinine (mg/dl) | 0.65 | 0.25 |
| **Liver function test** |  |  |
| Alanine transaminase (IU/L) | 20.67 | 0.99 |
| Aspartate transaminase (IU/L) | 21.82 | 8.61 |
| Alkaline phosphatase (IU/L) | 109.69 | 68.07 |

### Comparison of disease severity based on age and sex

A chi-square/ Fischer’s exact test was run to compare the presence of statistically significant difference based on age group and sex. Accordingly, no statistically significant difference was observed between the age groups and sex. (**Table 3**)

**Table 3:** Comparison of disease severity based on age and sex among children with COVID-19 (n=90)

| **Variables** | **Disease severity** |  |  | **Total (%)** | **P-value** |
| --- | --- | --- | --- | --- | --- |
|  | **Asymptomatic (%)** | **Mild (%)** | **Moderate (%)** |  |  |
| **Age group (in** |  |  |  |  |  |
| **years)** |  |  |  |  |  |
| ≤ 10 | 21 (35.6) | 3 (21.4) | 2 (11.8) | 26 (28.9) | 0.061 |
| 11-15 | 20 (33.9) | 7 (50.0) | 4 (23.5) | 31 (34.4) |  |
| 15-18 | 18 (30.5) | 4 (28.6) | 11 (64.7) | 33 (36.7) |  |
| **Sex** |  |  |  |  |  |
| Female | 36 (61.0) | 9 (64.3) | 12 (70.6) | 57 (63.3) | 0.854 |
| Male | 23 (39.0) | 5 (35.7) | 5 (29.4) | 33 (36.7) |  |

## DISCUSSION

In this study we have assessed the characteristics and outcome profile of 90 children with RT-PCR confirmed COVID-19 who were admitted to Millennium COVID-19 Care Center in Ethiopia from June 23 to September 17, 2020. Understanding this will guide the existing practice.

The median age of the participants was 15 (IQR, 10-17) years. A similar finding was observed in the North American study [4]. Two third of the participants were 11 to 18 years of age. This age group was also reported to have a higher disease incidence than the younger groups in a pattern that is recorded in the United States [5].

Almost two third of the participants were females unlike the pediatric studies conducted in other countries where the majority of infected children are males [3, 4]. In addition, researches conducted among adults in our center also showed a contrasting pattern where the majority of infected are males [19-24].

Close contact with a diagnosed person constituted the major (41/90) route of disease transmission. This was found to be the case in other studies as well where 66% up to 95.6% of admitted children were reported to acquire the disease through household contact with adults [6, 7]. This shows that there is a potential for an increase in the number of new infection among children since both globally and nationally the number of new cases among adults is increasing at an alarming rate from day to day implying the need for taking effective precautions among adults in order to protect the children.

Only three had a history of pre-existing comorbid illness, two had bronchial asthma and one had a history of valvular heart disease. This seems to be in contrast to other studies where 42% up to 83% of admitted children had a history of one or more pre-existing co-morbid illness [3, 4].

The majority (59/90) of the participants were asymptomatic at diagnosis. On the other hand, other studies reported 73% up to 94.1 % of children presenting with one or more symptom [4, 6, 8]. The most common symptom was cough, followed by sore throat, headache, fever, runny nose, chest pain, fatigue and nausea and vomiting. Cough was also reported to be the major presentation in another study in China [7] but fever was found to be the commonest presenting symptom in a number of studies including systematic review and meta-analysis studies that included more than 120 articles [6, 8, 9].

Among the entire pediatric admission during the study period in this largest COVID-19 Care center in the country, the majority (59/90) of the children were asymptomatic, 14 had mild disease and the rest 17 had moderate disease. There was no severe or critical case. On the contrary, studies from other countries revealed that a considerable proportion of children were diagnosed with severe and critical illness with a need of Intensive care unit admission and invasive ventilation [3-6, 8]. In addition, in the current study, there was no death among the study population. This is in contrast to a study conducted in United Kingdom and North America where mortality rate of 1% and 4% were reported, respectively [4]. This also shows that the disease might have a different pattern compared to adults as a study conducted at our Center among adults reported a mortality rate of 5.3% [23]. This difference could be attributed to the large proportion of asymptomatic patients in the studied population and also the small sample size in our study.

The variation in report among studies could be attributed to the small sample sizes included in most studies or due to the difference in underlying population characteristics and difference in admission criteria between countries. Therefore, though the finding from this study can be used as a baseline, further study with large sample size is necessary to have a better understanding of the pandemic in pediatric group as the small sample size population included in this study might not generalize the actual pattern and outcome of the disease at the national level.

## CONCLUSION

The majority of the identified cases were asymptomatic and there was no death during the observation period. The pediatric patients seemed to have a milder disease presentation and favorable outcome compared to other countries report and also the adult pattern observed in our country. Further multicenter study with a large sample size is recommended to gain a better perspective of the disease in children.

### What is already know on this topic

– COVID-19 among children is reported to have a diverse outcome among different countries with majority reporting a favorable disease course and outcome than adults.
– There is scarcity of research among children with COVID-19 compared to the adults, especially in the African set up.

### What this study adds

– This study assessed children with COVID-19, it is the first of few studies in Africa
– Pediatric patients seemed to have a milder disease presentation and a favorable outcome compared to the adult pattern observed in our country and other set ups.
– The finding can be used as a baseline for further large scale study and can be used to guide the practice in similar the mean time

## Data Availability

All relevant data are available upon reasonable request.

## Declaration

## Acknowledgment

The authors would like to thank St. Paul’s Hospital Millennium Medical College for facilitating the research work.

## Authors Contribution

TWL conceived and designed the study, revised data extraction sheet, performed statistical analysis, and drafted the initial manuscript. ISH, EHM, WCZ, NWC, MGE, DSA and MAA: contributed to the conception, obtained patient data, undertook review and interpretation of the data, revised the manuscript and approved the final version

## Availability of data and materials

All relevant data are available upon reasonable request.

